# Mapping the global research landscape of Nipah Virus (NiV): A bibliometric analysis of trends and collaborations

**DOI:** 10.1101/2025.03.29.25324876

**Authors:** Ashutosh Kumar Maurya, Ashish Kumar, Sristhi pandey, Deepak Verma, V.B. Sameer Kumar, Rajendra Pilankatta

## Abstract

Infectious diseases, also known as communicable disease or transmissible disease is illness that is caused due to pathogens such as bacteria, fungi, parasites and viruses. One of such infection is caused by zoonotic virus (bat-borne) known as Nipah virus (NiV). NiV is one of the world’s deadliest viruses leading to highest mortality rates. It is a single-stranded RNA virus belonging to category C priority pathogen that belongs to genus Hepanivirus, and family Paramyxoviridae. The first occurrence of NiV was observed in Malaysia-Singapore during 1998-1999 and the most recent outbreak in Kozhikode district of Kerala in 2018.

Currently, no drugs or vaccines specific to NiV have proven to yield effective outcome at advanced stage of the disease. However, some therapies involving antiviral drug like Ribavirin along with other medicines have shown to recover patients, reducing the death rate by 36%. Due to extensive research various therapeutic strategies have been proven to successfully target the viral infection during the early stages in animal models. Several vaccines have been under investigation and clinical trials serving as a promising approach towards effective and safe vaccine in both livestock animals and humans.

This study underscores the essential contributions shaping the academic discourse in nipah virus research, bringing to light the most influential studies within the co-citation and co-occurrence network analysis. By identifying the keywords, we gain valuable insights into the foundational research driving advancements in this research area. The detailed mapping of the co-citation network further illuminates the interconnectedness of various research themes and the collaborative efforts that boost the field of nipah virus research in different horizons. The recognition of influential studies not only highlights the depth and breadth of scholarly engagement but also provides a roadmap for future research endeavours.

## Introduction

Infectious diseases also known as communicable disease or transmissible disease is illness that is caused due to pathogens such as bacteria, fungi, parasites and viruses when they invade the host and multiply within them. One of such infection is caused by zoonotic virus (bat-borne) known as Nipah virus (NiV)[1].

NiV is one of the world’s deadliest viruses leading to highest mortality rates[2]. It is a single-stranded RNA virus belonging to category C priority pathogen that belongs to genus Hepanivirus, and family Paramyxoviridae [3]. The first occurence of NiV was observed in Malaysia-Singapore during 1998-1999. However, when the first case was reported from Kampung Sungai Nipah town of Malaysia, during an outbreak among pig farmers in the year 1999, it got its name as “Nipah Virus”[4]. Since then, many outbreaks have been reported in countries of South-east Asia such as in Singapore, Philippines, India and Bangladesh[5]. In India, the first outbreak happened in Siliguri and Nadia district of West Bengal in 2001 and 2007 respectively, following the most recent outbreak in Kozhikode district of Kerala in 2018[6-8]. Out of all the total incident cases reported worldwide, Bangladesh and India contributed 42% and 15% cases respectively whereas Malaysia accounted for 43% cases during the outbreak.

Fruit bats belonging to Pteropus genus are considered as the primary reservoir host of this emerging highly pathogenic zoonotic virus found in West Africa and Madagascar, as well as south-east Asia[9]. It is not only confined to humans but also possess the ability to infect wide range of animals including pigs, monkeys, horses and other domestic animals such as goats, sheep, cat and dogs[10-11]. In pigs, the disease is also known as porcine respiratory and encephalitis syndrome (PRES) also known as barking pig syndrome (BPS) [12]. The pigs may serve as an intermediate symptomatic host transmitting the disease to humans. One of the key routes of transmission of NiV involves either direct or indirect contact with infected secretions or bodily fluids of animals like saliva, urine and excreta such as in the case of southern state, Kerala where the person was affected on consumption of fruits contaminated by NiV infected bats[13]. In Malaysia the people were infected with the disease from NiV infected pigs [14]. Import of NiV infected pigs to Singapore from Malaysia led to the spread of infection to 11 pig farmers with one case of death [15]. The other way involves direct human-to-human transfer such as in the case India (West Bengal and Kerala) and Bangladesh where epidemiologic similarities were observed among the people who were contracted from NiV during the healthcare settings while giving treatments to the NiV infected patients which could possibly be due to coming in contact with patient’s bodily fluids like blood, saliva, urine including respiratory and pharyngeal secretions [16-17]. Along with the factors mentioned above, the other possible causes of NiV outbreak are due to environmental, ecological and anthropogenic influences such as climate change, deforestation, industrialization and farming leading to a drastic disruption of the ecosystem and environment [18-19].

NiV gains entry inside the targeted host via micropinocytosis leading to initial symptoms of vomiting, fever, headache, cough, muscle aches and sore throat which is followed by altered consciousness, dizziness, respiratory disorders such as acute respiratory distress syndrome(ARDS) and some neurological signs indicating encephalitis (brain swelling) resulting in multiple organ dysfunction syndrome (MODS) leading to coma and death in severe cases[20]. The incubation period of infected patients after exposure to NiV ranges from few days on an average of < 2 weeks as seen in case of Kerala and Bangladesh outbreak and can extend up to 2 months as observed during Malaysian outbreak[12,22-23]. The fatality rate of the disease has been statistically estimated to be 40-75% depending upon the encephalitic and severity of the case.

In the initial stages of NiV infection the signs and symptoms are non-specific hindering the accuracy of the diagnosis and pose a challenge for effective control measures. However, diagnosis can be performed on the patients with a clinical history or during their recovery period. Among all the diagnostic tests available, PCR including RT-PCR, nested PCR and qRT-PCR are the most preferred due to its high sensitivity and capability to detect rapid mutation virus has undergone by targeting its conserved segments M, N and P of genome. [24-25]. Another alternative approach is a serological assay, ELISA which aids in detection of viral antigen and antibody response. The confirmatory tests include viral isolation, neutralization and histopathology in some fatal cases [26-27]. In humans the preferred samples specimens are throat swap, urine, blood, cerebrospinal fluid whereas for the isolation of NiV, tissue samples from the organs of dead animals are preferred [28]. All these diagnostics methods are restricted to BSL-4 facilities.

Currently, no drugs or vaccines specific to NiV have proven to yield effective outcome. However, some therapies involving antiviral drug like Ribavirin along with other medicines have shown to recover patients, reducing the death rate by 36% whereas in the case of animals Favipiravir and monoclonal antibodies have proven its efficacy[29-30]. Recent studies have found that NiV remains viable in sugary pulp of fruits for several days and can be transmitted upon its consumption. Owing to unavailability of proper vaccination strategy till now, adopting various preventive measures can help prevent transmission of disease. Outbreaks of SARSCoV2 and Ebola virus have led to the implementation of guidelines to protect the healthcare workers. Increasing awareness and following of strict protocols formed by national healthcare authorities can help minimize the contact of healthcare workers with the infected individual during patient’s care[31-32]. Other preventive techniques involve vaccination of the animal livestock and keeping the grazing lands and animal farms away from the fruit trees as well as bat roosting trees because they can act as an intermediate host for spreading the disease [33]. Surveillance tool can also possibly serve as an early detection tool in patients and livestock to help prevention [34].

Due to extensive research various therapeutic strategies have been proven to successfully target the viral infection during the early stages in animal models. Several vaccines have been under investigation and clinical trials serving as a promising approach towards effective and safe vaccine in both livestock animals and humans. Additionally, the goal is to increase the rate of clinical test and improve the quality of molecular assays contributing to early detection of virus with accuracy.

## Methodology

This section delves into the pivotal initial phase of our investigation: curating a targeted collection of documents essential to addressing the research objectives. A rigorous and methodologically sound analysis hinges on the strategic selection of high-quality, relevant sources. Here, we elucidate the structured approach adopted to identify, evaluate, and prioritize literature and data directly tied to understanding the Nipah virus outbreak. We delineate the inclusion criteria, search strategies, and validation processes designed to ensure alignment with the scope and questions of the present study. By transparently outlining this systematic compilations process, we lay the groundwork for an authoritative, evidence-driven exploration of the outbreak’s dynamics, challenges, and implications.This version sharpens the focus on the Nipah virus context while emphasizing methodological rigor, transparency, and relevance to public health research.

### Identifying sources for the review

The Scopus-indexed database was utilized to extract and retrieve documents relevant to the Nipah virus, capitalizing on its recognized utility in Bibliometric and public health research (Singh et al., 2021). Prior studies, including those by Singh et al.(2021)Singh et al. (2021), underscore(Singh et al., 2021) interdisciplinary breadth of Scopus database, particularly in biomedical and epidemiological fields, where it offers broader coverage compared to alternatives databases like Web of Science. For instance, in virology, outbreak management, and zoonotic disease research domains critical to understanding the transmission dynamics of Nipah virus, pathogenesis, and containment strategies. The extensive indexing of Scopus database ensures access to diverse scholarly outputs, ranging from clinical studies to regional outbreak reports (Singh et al., 2021).

Critics posit that databases such as Web of Science and Dimensions prioritize selectivity, emphasizing high-impact, peer-reviewed journals, which may enhance source quality but has limited scope (Hallinger & Kovačević, 2019; Singh et al., 2021). However, the authors argue that database suitability is context-dependent, necessitating empirical validation within specific research frameworks. In the case of the Nipah virus a pathogen requiring interdisciplinary insights from virology, epidemiology, ecology, and public health the inclusivity of Scopus proves advantageous for capturing fragmented or regionally dispersed studies, such as outbreak investigations in South and Southeast Asia (Singh et al., 2021). To address potential biases, supplementary validation through manual screening of sources, including preprints and regional databases (e.g., India’s ICMR repositories), was implemented to ensure rigor and relevance.

By employing Scopus database, this study balances comprehensive coverage with methodological credibility, a vital consideration for Bibliometric analyses aiming to map global research trends, therapeutic developments, and policy responses to Nipah virus outbreaks (Hallinger & Kovačević, 2019; Singh et al., 2021).Given the limited availability of consolidated data on Nipah virus transmission dynamics and outbreak patterns, researchers retrieved 754 documents through a targeted search strategy. The search query *“Nipah AND virus AND human”* was applied to the Article title, Abstract, and Keywords fields to capture studies addressing virological, epidemiological, and public health dimensions of the pathogen (Singh et al., 2021). To mitigate potential gaps, supplementary manual searches of grey literature and public health repositories (e.g., WHO Outbreak News) were conducted, though the core dataset remains Scopus-derived to maintain Bibliometric severity.

This analysis focuses on literature published between 2000 and early February 2025. It specifically examines research that includes the terms “Nipah” and “Virus” and “human” across a wide range of subject areas, such as Immunology and Microbiology, Biochemistry, Genetics and Molecular Biology, Agricultural and Biological Science, Multidisciplinary, Pharmacology, Toxicology and Pharmaceutics, Veterinary, Neuroscience, Environmental Science, Arts and Humanities, Social Science, Nursing and Psychology. The current study encompasses all document types, excluding Erratum, due to concerns regarding its reliability and authenticity. Any document types which were not written in English were excluded from the study to ensure consistency in language and to avoid potential inaccuracies or misinterpretations. This approach helps maintain the integrity and clarity of the analysis.

To ensure a transparent and systematic search process, this review followed the Preferred Reporting Items for Systematic Reviews and Meta-Analyses (PRISMA) guidelines (Moher et al., 2009). The search term “Nipah Virus and human” was used as a query in the Scopus database, resulting in an initial retrieval of 756 documents. After applying exclusion criteria such as removing retracted articles, documents lacking author or index keywords, and those not in English, 29 documents were eliminated. The Bibliometric data was extracted via CSV files on 10^th^ February 2025, culminating in a final database of 725 documents. This collection includes Conference Papers, Articles, Reviews, Book Chapters, Notes, Editorial, Short Survey and Letter.

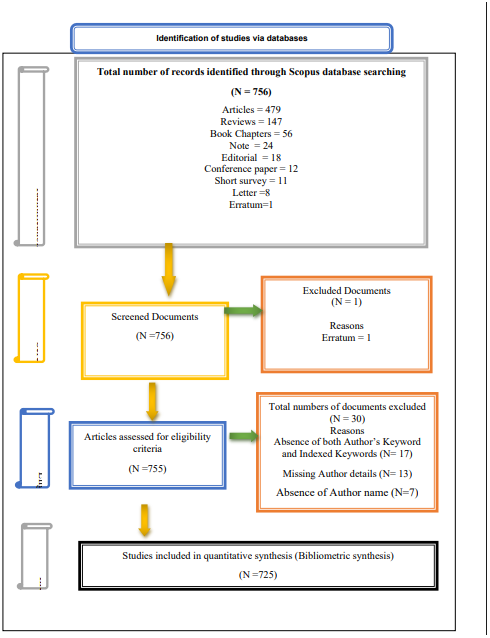

**PRISMA flow diagram for the identification of documents.** (Preferred Reporting Items for Systematic Reviews and Meta-Analyses (PRISMA) flow diagram elaborated steps in the identification and screening of data sources)

### Data Analysis

For the Bibliometric analysis, detailed bibliographic information from the 725 documents such as authors, publication years, titles, keywords, affiliations, citations, and related metadata was exported and systematically organized. The analysis employed descriptive statistics alongside advanced Bibliometric techniques, including citation analysis, co-citation analysis, author co-citation analysis, and keyword co-occurrence analysis, to facilitate the “visualization of similarities” (Small, 1999; van Eck & Waltman, 2014; White & McCain, 1998; Zupic & Čater, 2015). The Bibliometric studies were conducted using VOSviewer software (van Eck & Waltman, 2014), supplemented by Excel and Scopus analytical tools.

## Results

### Descriptive analysis of publication on NIPAH virus

#### 1. Allocation of yearly publication trends

In the present study, bibliometric methods were used to map the global research on Nipah virus with particular emphasis on understanding the molecular mechanism of infection and drug development. The first case of nipah virus outbreak occurred in 1999 and a total of 725 publications were released between 2000 and February 2025. As per yearly publication information provided in table 1, a total of 9 publications came in the year 2000 followed by 3 in the year 2001 and 8 in 2002. From 2003 onwards a gradual increase in the publications was seen, where the number of publications ranged between 13 to 41 till the end of 2019. The number of publications jumped to 56 in the year 2020 followed by 47 and 38 in the year 2021 and 2022 respectively. In 2023 again the number of publications reached its peak and it further declined in 2024. Till the early 2025, a total of 8 publications were published and this number is still counting.

**Table :**
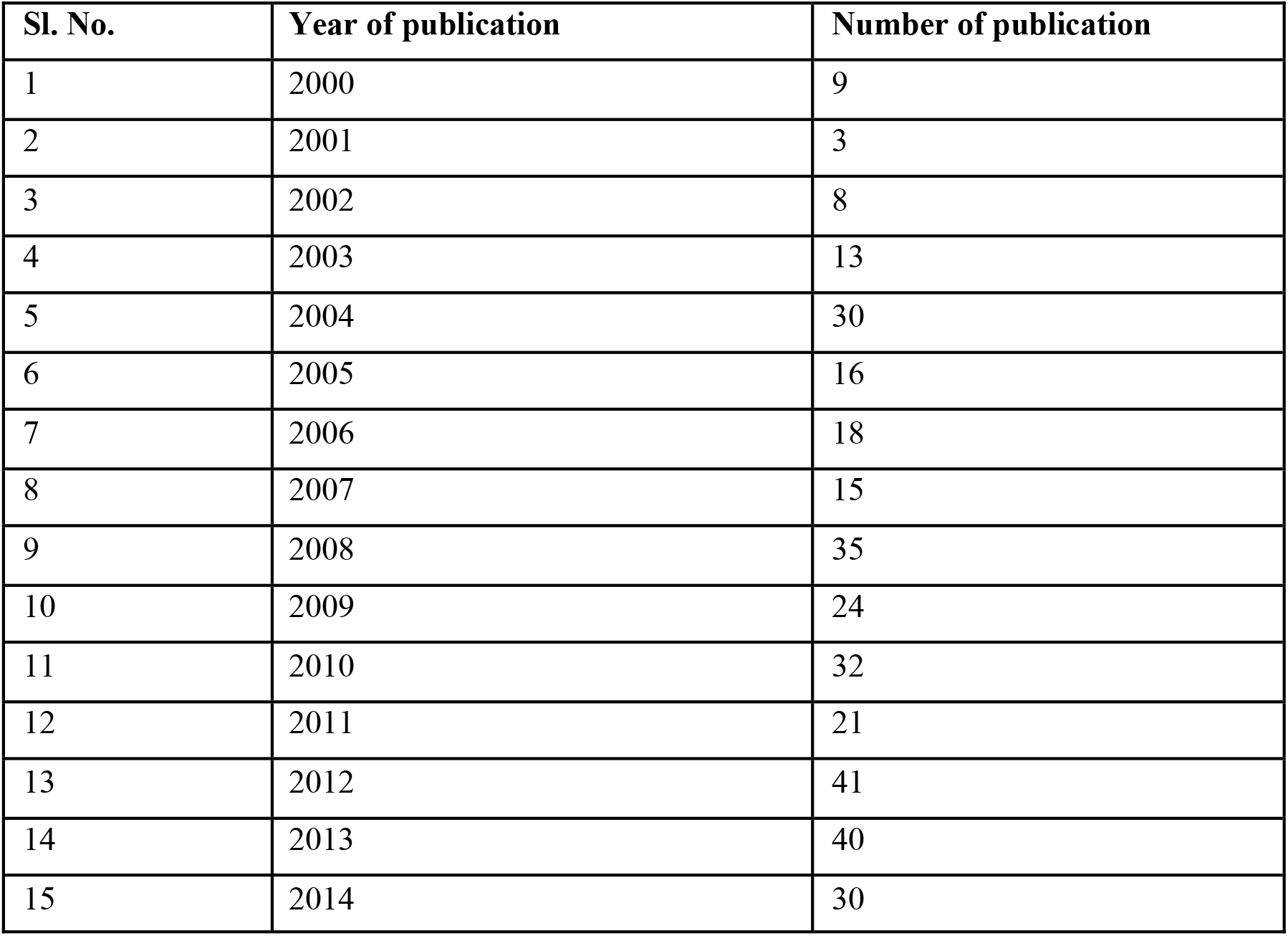
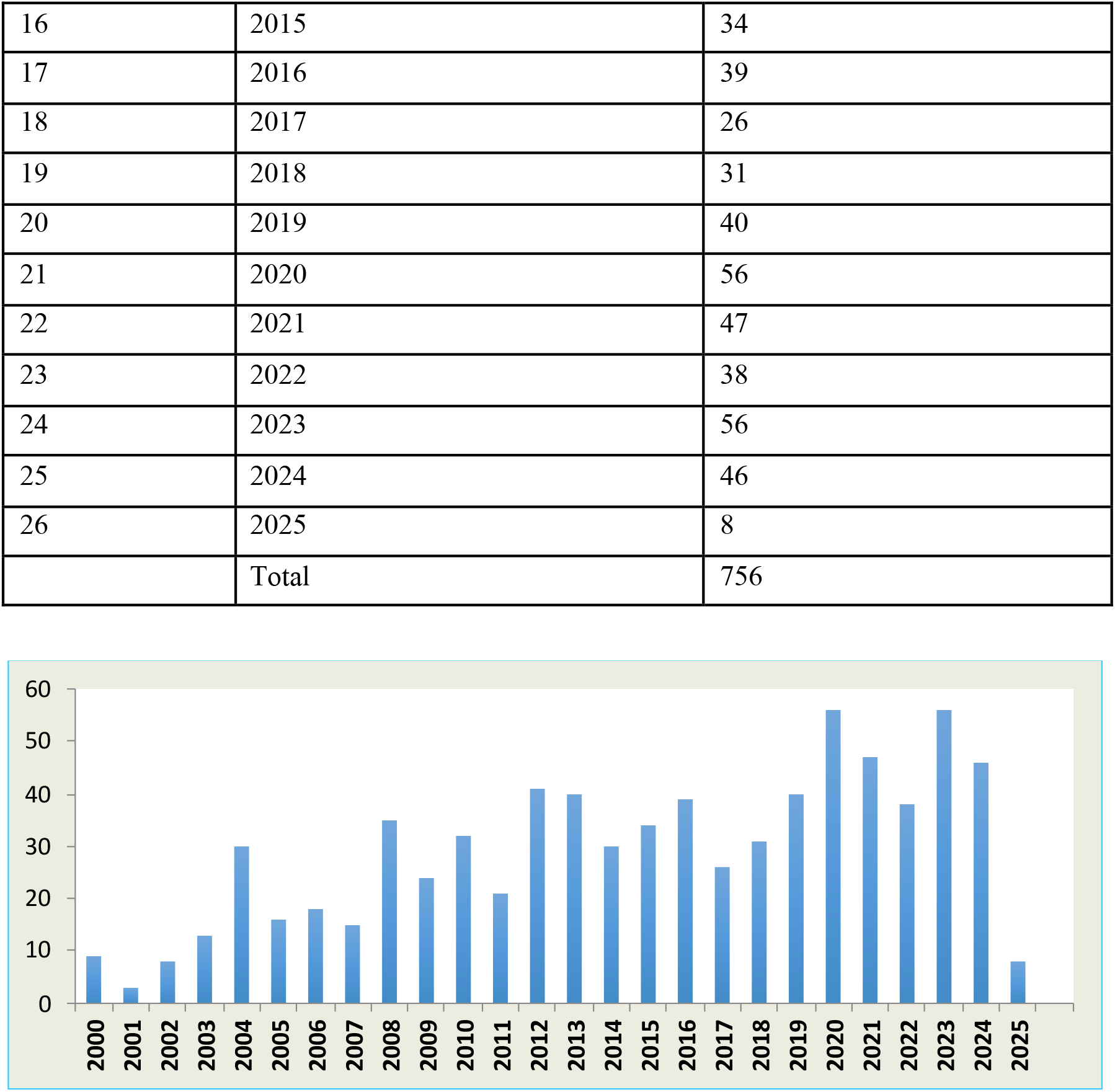
Allocation of yearly publication trends.

| Sl. No. | Year of publication | Number of publication |
| --- | --- | --- |
| 1 | 2000 | 9 |
| 2 | 2001 | 3 |
| 3 | 2002 | 8 |
| 4 | 2003 | 13 |
| 5 | 2004 | 30 |
| 6 | 2005 | 16 |
| 7 | 2006 | 18 |
| 8 | 2007 | 15 |
| 9 | 2008 | 35 |
| 10 | 2009 | 24 |
| 11 | 2010 | 32 |
| 12 | 2011 | 21 |
| 13 | 2012 | 41 |
| 14 | 2013 | 40 |
| 15 | 2014 | 30 |
| 16 | 2015 | 34 |
| 17 | 2016 | 39 |
| 18 | 2017 | 26 |
| 19 | 2018 | 31 |
| 20 | 2019 | 40 |
| 21 | 2020 | 56 |
| 22 | 2021 | 47 |
| 23 | 2022 | 38 |
| 24 | 2023 | 56 |
| 25 | 2024 | 46 |
| 26 | 2025 | 8 |
|  | Total | 756 |

#### 2. Distribution of publication by country

This section deals with the countries having highest number of publications representing the research on nipah virus. Table 2 shows the top 10 countries having highest number of publications from the year 2000 to February 2025, in descending order. With our analysis, we found that the United states published maximum number of articles (396) followed by Australlia (130) and India (73) respectively. Further, United kingdom published 68 documents, Geramany (56), France (52), Canada (39), China (35), Singapore (29) and Bangladesh (26). Our list also contains 31 publications from unidentified sources.

**Table 2:** Country-wise distribution of publications.

| Sl. No. | Country | No. of documents |
| --- | --- | --- |
| 1 | United States | 396 |
| 2 | Australia | 130 |
| 3 | India | 73 |
| 4 | United kingdom | 68 |
| 5 | Germany | 56 |
| 6 | France | 52 |
| 7 | Canada | 39 |
| 8 | China | 35 |
| 9 | Undefined | 31 |
| 10 | Singapore | 29 |
| 11 | Bangladesh | 26 |

#### 3. Distribution of articles published by subject area

This sections represents the number of publications, came from various disciplines from 2000 to February 2025 on nipah virus. As evidenced from the table 3, highest number of publications came from Immunology and microbiology research area (345), followed by 223 publications from the field of Biochemistry, Genetics and Molecular Bilogy. Further, we found that a total of 172 publications came from Agriculture and Biological science research, followed by 127 publications was reported from multidisciplinary areas. Along with this, 64 publications came from Pharmacology, Toxicology & Pharmaceutics, 54 documents were published from veterinary science, 25 reports from Neuroscience, 25 from Environmental science, 10 from Arts & Humanities, 6 from Social science, 2 from Nursing and 1 from the Psychology discipline.

**Table 3:**
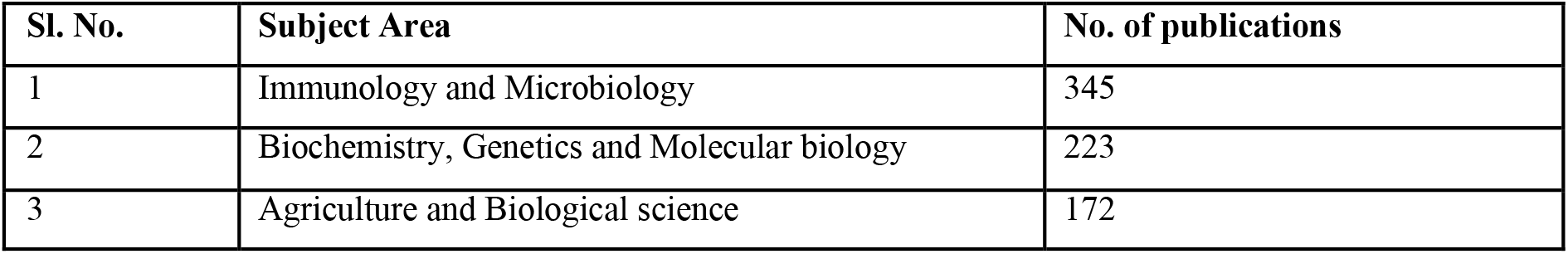
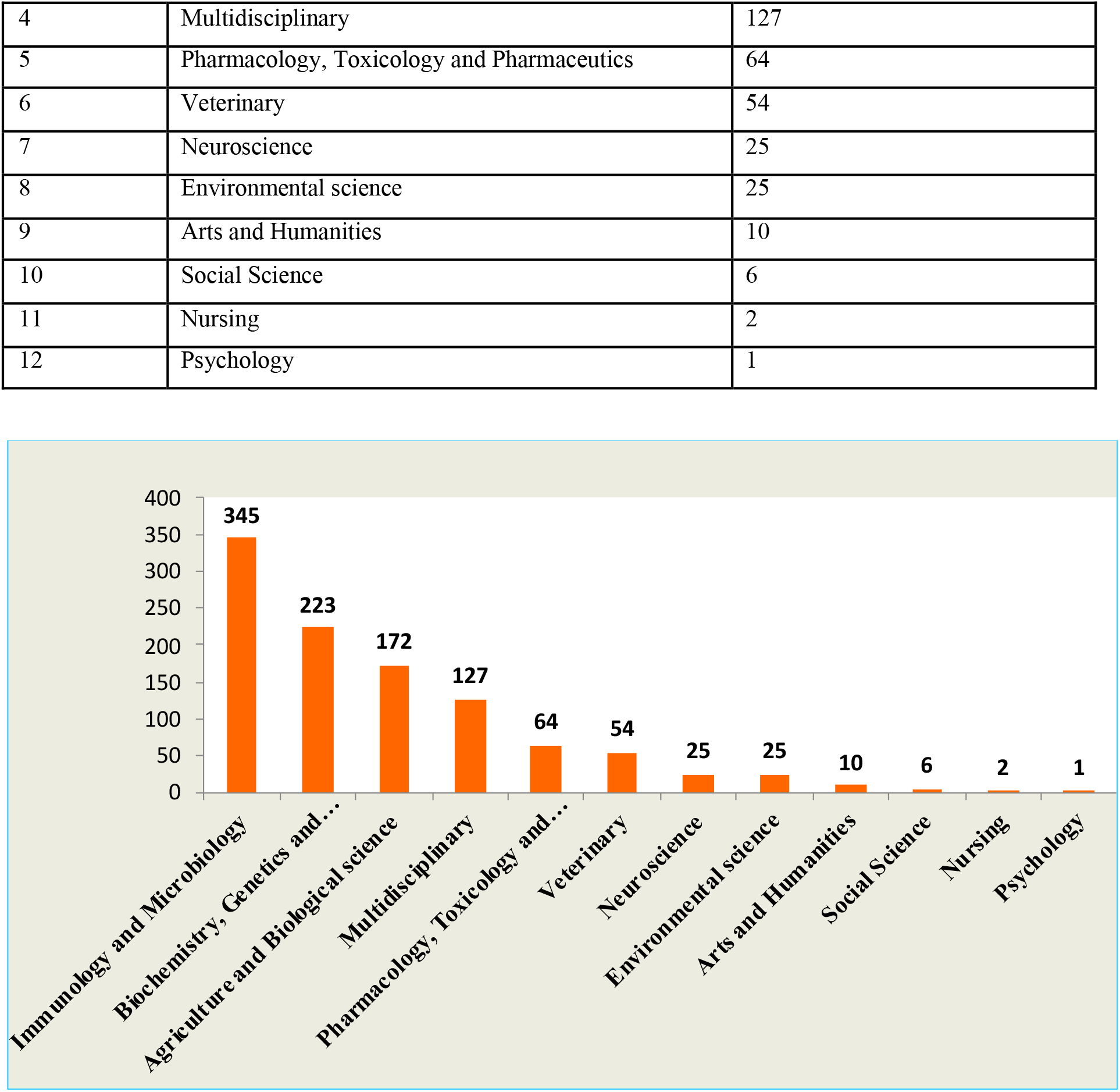
Subject area wise distribution of publications.

| Sl. No. | Subject Area | No. of publications |
| --- | --- | --- |
| 1 | Immunology and Microbiology | 345 |
| 2 | Biochemistry, Genetics and Molecular biology | 223 |
| 3 | Agriculture and Biological science | 172 |
| 4 | Multidisciplinary | 127 |
| 5 | Pharmacology, Toxicology and Pharmaceutics | 64 |
| 6 | Veterinary | 54 |
| 7 | Neuroscience | 25 |
| 8 | Environmental science | 25 |
| 9 | Arts and Humanities | 10 |
| 10 | Social Science | 6 |
| 11 | Nursing | 2 |
| 12 | Psychology | 1 |

#### 4. Distribution based on document type

In this study, we selected 725 publications falling in 8 different class of documents, which has been summarised in Table 4. Our analysis revealed that research articles contribute the maximum number of publications with 479 documents followed by 147 review articles on nipah virus. Apart from research and review articles, a considerable number of documents were published as book chapter (56), Note (24), Editorial (18), Conference paper (18), Short survey (11) and Letter (8). In our study we have excluded Erratum (1) document, identified at initial stage.

**Table 4:** Document wise distribution of publication.

| Sl. No. | Document type | No. of Publications |
| --- | --- | --- |
| 1 | Article | 479 |
| 2 | Review | 147 |
| 3 | Book chapter | 56 |
| 4 | Note | 24 |
| 5 | Editorial | 18 |
| 6 | Conference paper | 12 |
| 7 | Short survey | 11 |
| 8 | Letter | 8 |

#### 5. Analysis of influential authors and documents

In this section, we have listed the top 10 most influential authors based on the citations on their work in the field of Nipah virus research by setting the filter at minimum 2 documents and minimum citations 300. Citation analysis is one of the common criteria in Bibliometric to assess the academic and scientific impact of a research work within a specific research domain. Table 5 shows the frequently cited author along with their research paper and journal details in descending order. The research work of Wang Lin-fa et al. have the highest number of citations (4437), followed by Broder C.C. et al. with 3079 citations. Further, the research work of Lee behnur et al. has 2298 citations, Cramery G. et al. 2205 citations, Bossart K.N et al. 1809 citations, Middleton D. et al. 1530, Aguilar H.C et al., 1496, Chua K.B et al. 1346, Basler C.F. et al 1239 and Dimitrov D.S. et. al 1205.

**Table 5:**
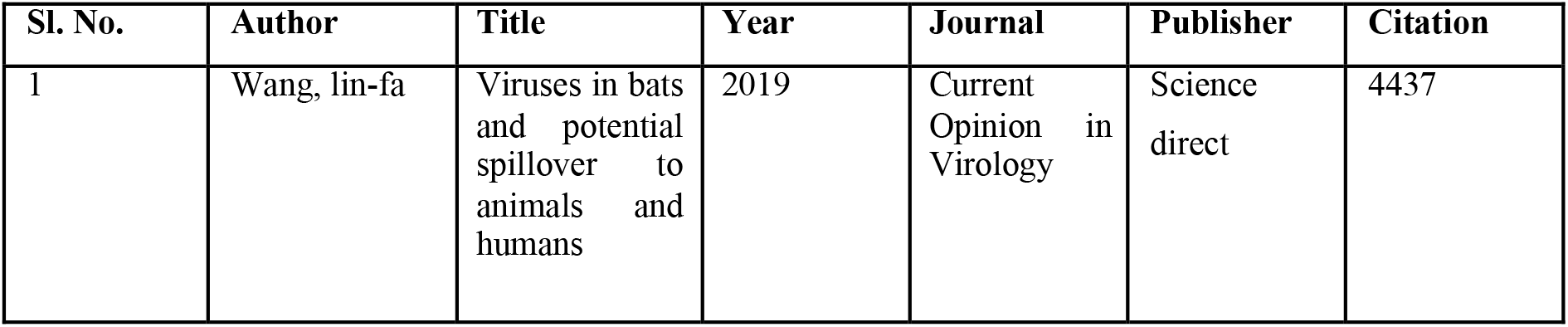
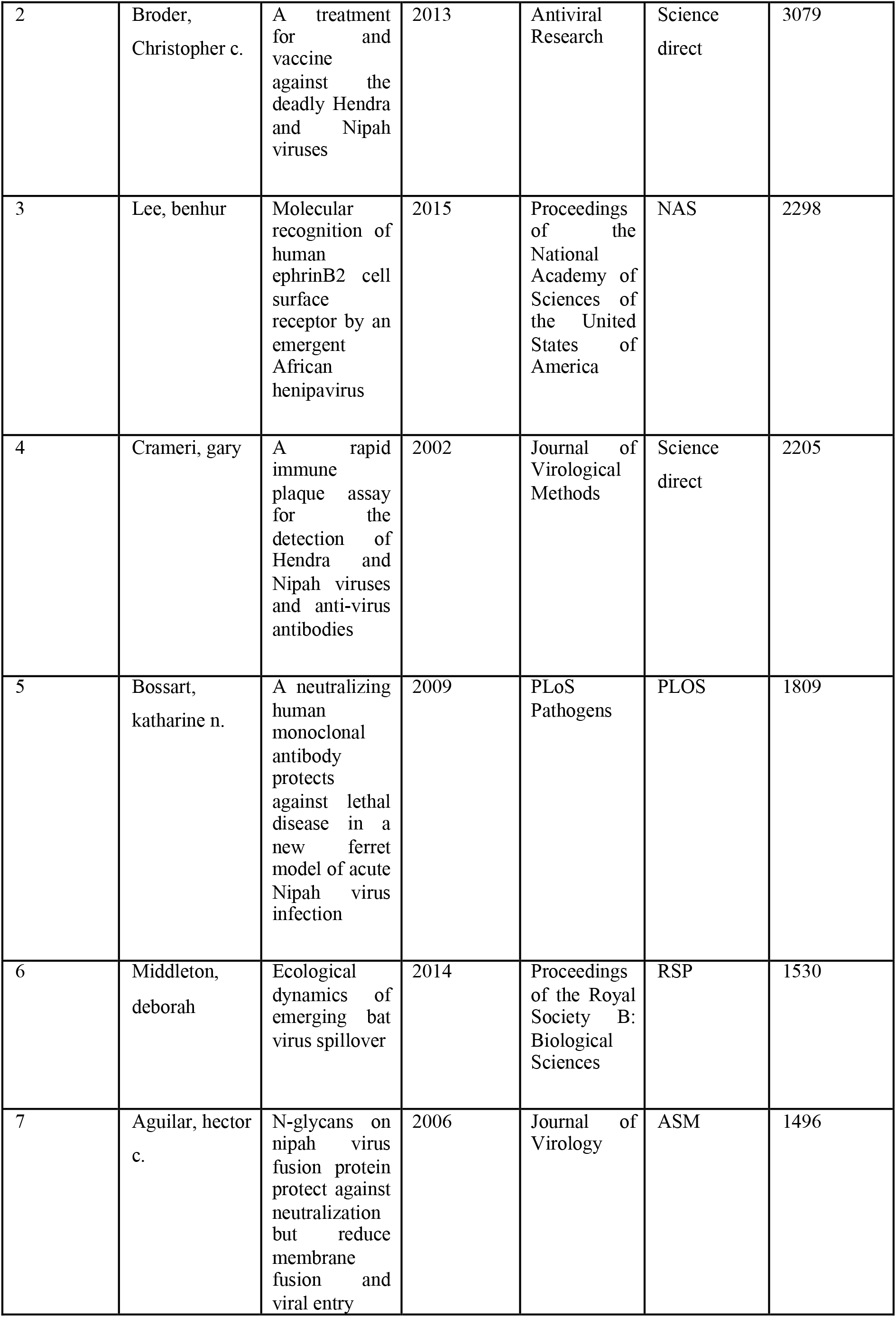
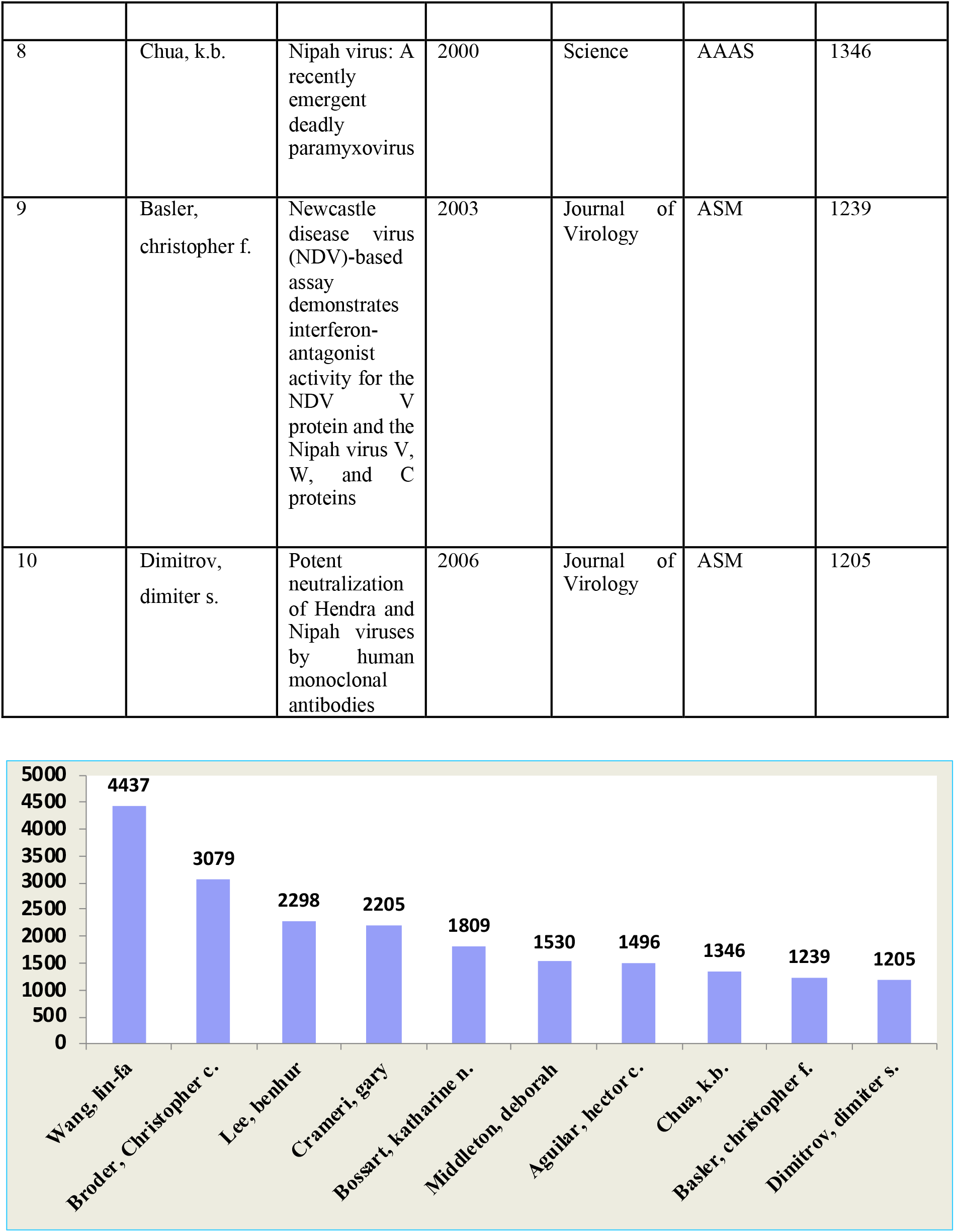
Distribution of influential authors and citation counts.

The information presented in table 5 provides the insights for the researchers to find the pioneer contributors in the field of nipah virus research along with highly impactful literature.

#### 6. Analysis of journal citations

This section focuses on the citation score of top 10 journals publishing the research work on by a journal as 10 and minimum number of citations as 50.

An extensive exploration of the distribution of publications across various journals revealed that the journal of virology scored the highest number of citations 7527, followed by Science journal with a citation of 2387. Following these, nature journal has a citation score of (2333), Plos One (2309), Plos Pathogen (1583), PNAS (1580), Antiviral research (1084), current opinion in virology (1046), Virology (1000) and Journal of general virology (928).

Further, table 6 also shows the quartile (Q-index) and Hirsch (h-index) values derived from Scimago journal & country rank for the top 10 journals, distinguished by highest number of significant publication. The h-index is an author level indicator that assesses the quality of research over time while taking into account the author’s scholarly output as well as the effect of their study. Nature journal has the highest h-index of 1391 followed by Science journal (1336), PNAS (869), Plos One (435), Journal of virology (321), Plos pathogens (246), Virology (195), Journal of general virology (187), Antiviral research (151) and Current opinion in virology (94). 8 out of 10 selected journals have Q1 quartile ranking where as 2 journal holds Q2 rank.

**Table 6:**
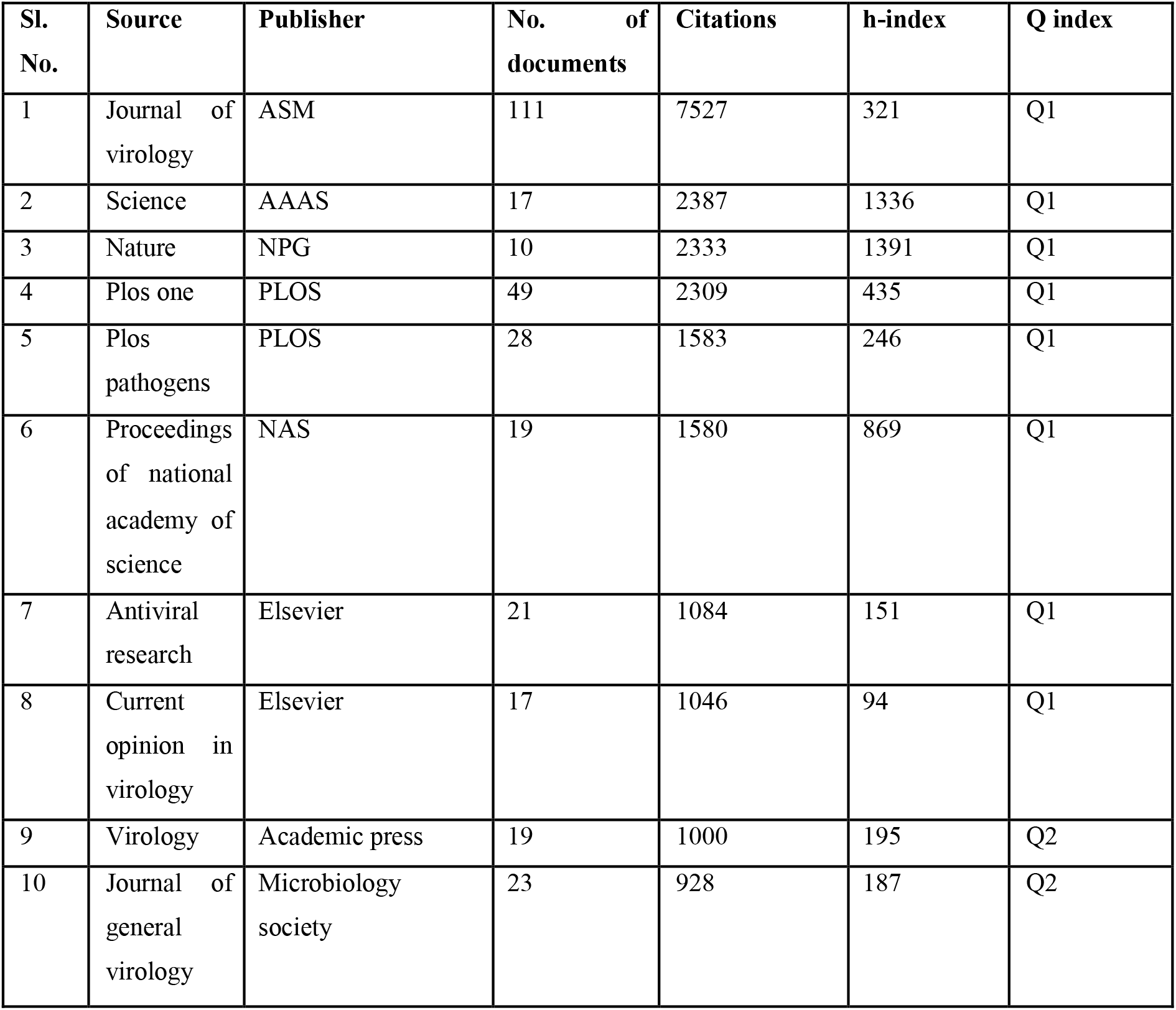
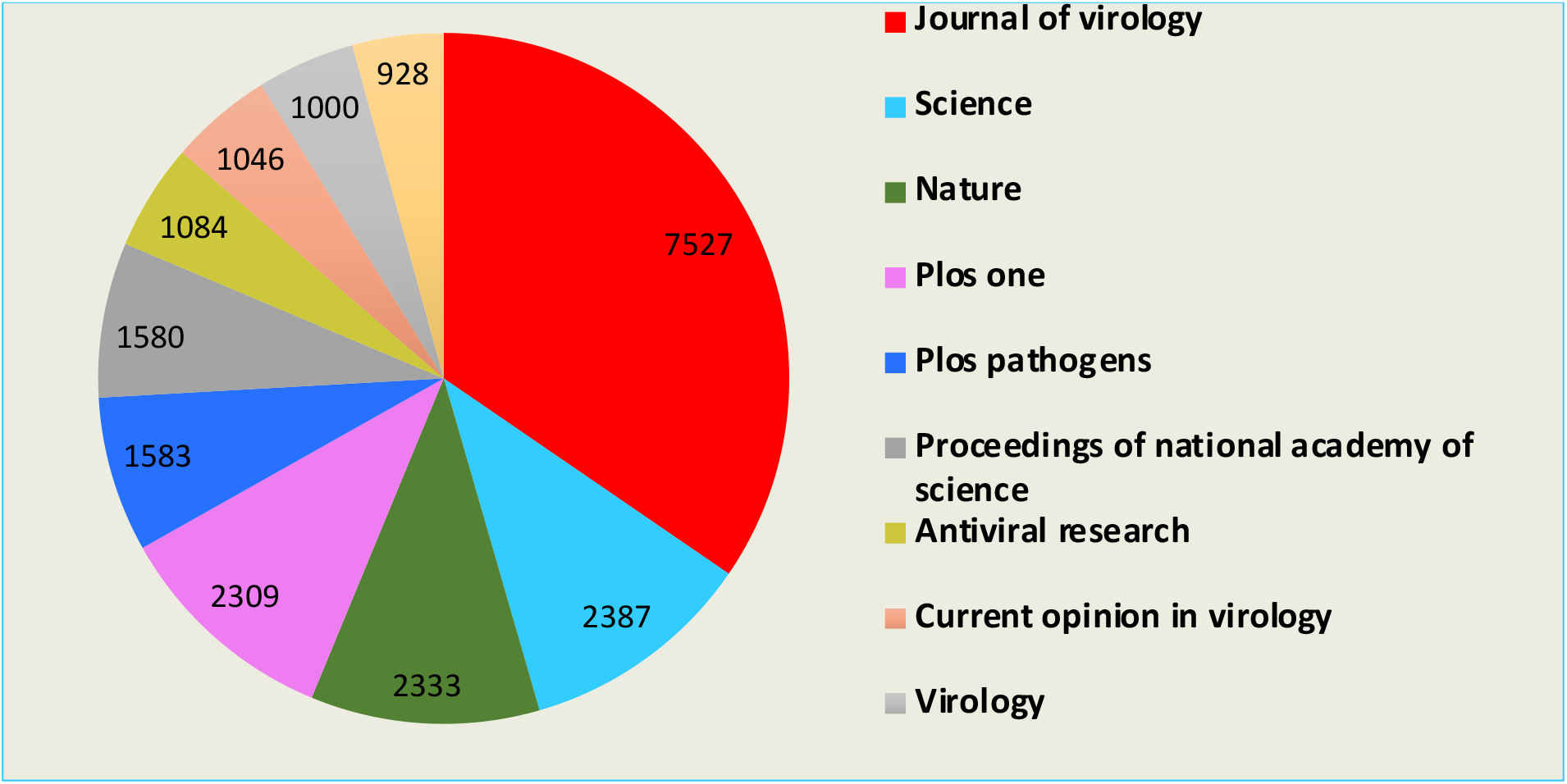
Distribution of journals with highest publications.

#### 7. Co-authorship exploration for countries

The co-authorship exploration of the research publications based on the countries, published in the scopus indexed journals between 2000 to 2025 is displayed in figure 1. A country is represented by a circular form (Node), larger circles indicate greater influence. The collaboration beween the 2 countries has been represented by a line (Link). The node distance and the link thickness shows, how closely the countries are collaborating.

**Figure 1:**
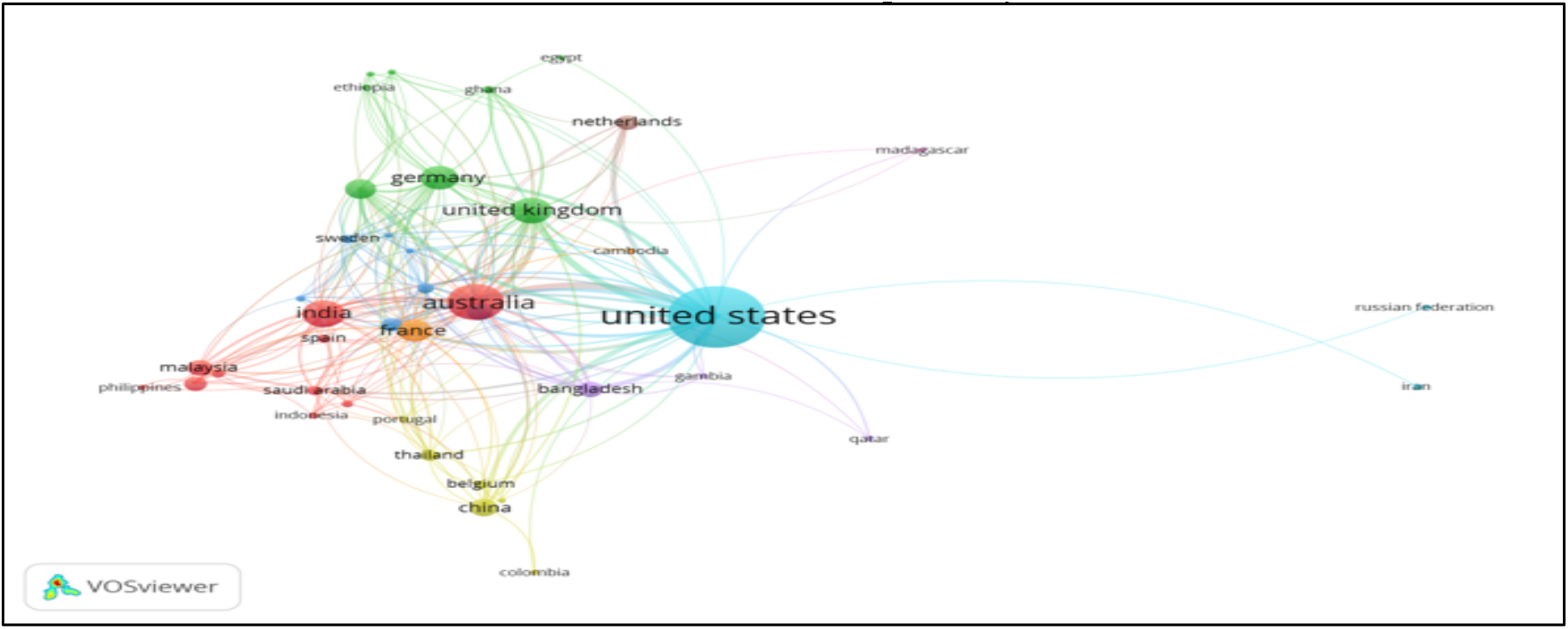
Co-authorship exploration by country.

The analysis of co-authorship revealed that out of 80 countries, only 25 meet the threshold of minimum 2 documents with 10 citations. The United States collaborated with maximum countries followed by Australia and India respectively.

#### 8. Co-authorship exploration for organisations

Figure 2 illustrates the network analysis of co-authorship among organizations. To achieve this, we have set a threshold requiring a minimum of 5 document and atleast 25 citations. Out of 2059 organizations meeting these criteria, a double cluster emerged, consisting of 17 interconnected organizations. This network highlights the collaborative dynamics within the field, emphasizing the importance of both document production and citation impact in forming significant research partnerships.

**Figure 2:**
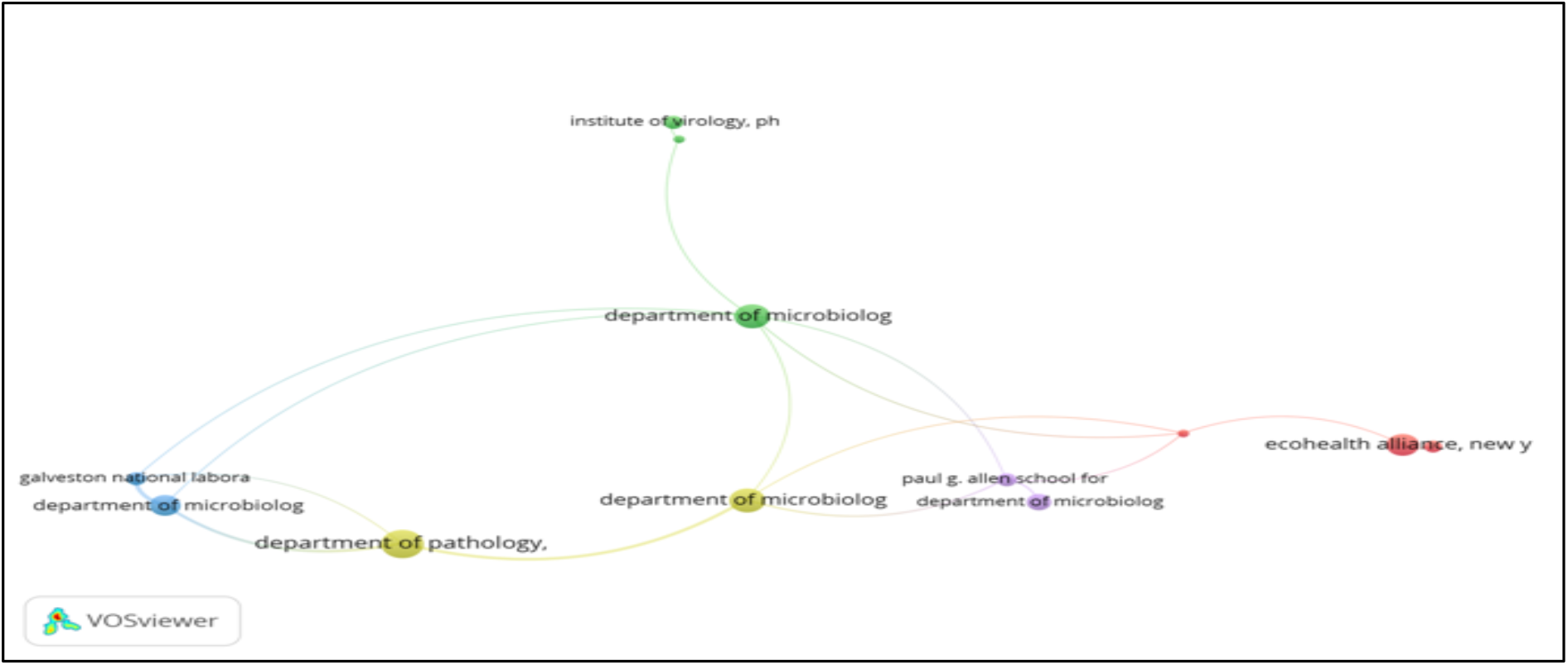
Co-authorship exploration by institution.

The results revealed that, the Department of pathology, University of Texas, USA (17 documents & 12 links) has the highest number of documents along with the highest number of collaborations with the other organizations. Department of microbiology and immunology, Uniformed Services University, USA occupies second position in terms of number of documents and total link strength.

#### 9. Co-authorship exploration for Authors

The co-authorship network among authors was analyzed using VoS Viewer software keeping minimum number of document published by an author as 1 having atleast 50 citations and has been represented in figure 3. Analysis revealed that the Morens et al. (1555) has the highest number of citations followed by Chua K.B (1064) and lloyd-smith et al. (499), respectively.

**Figure 3:**
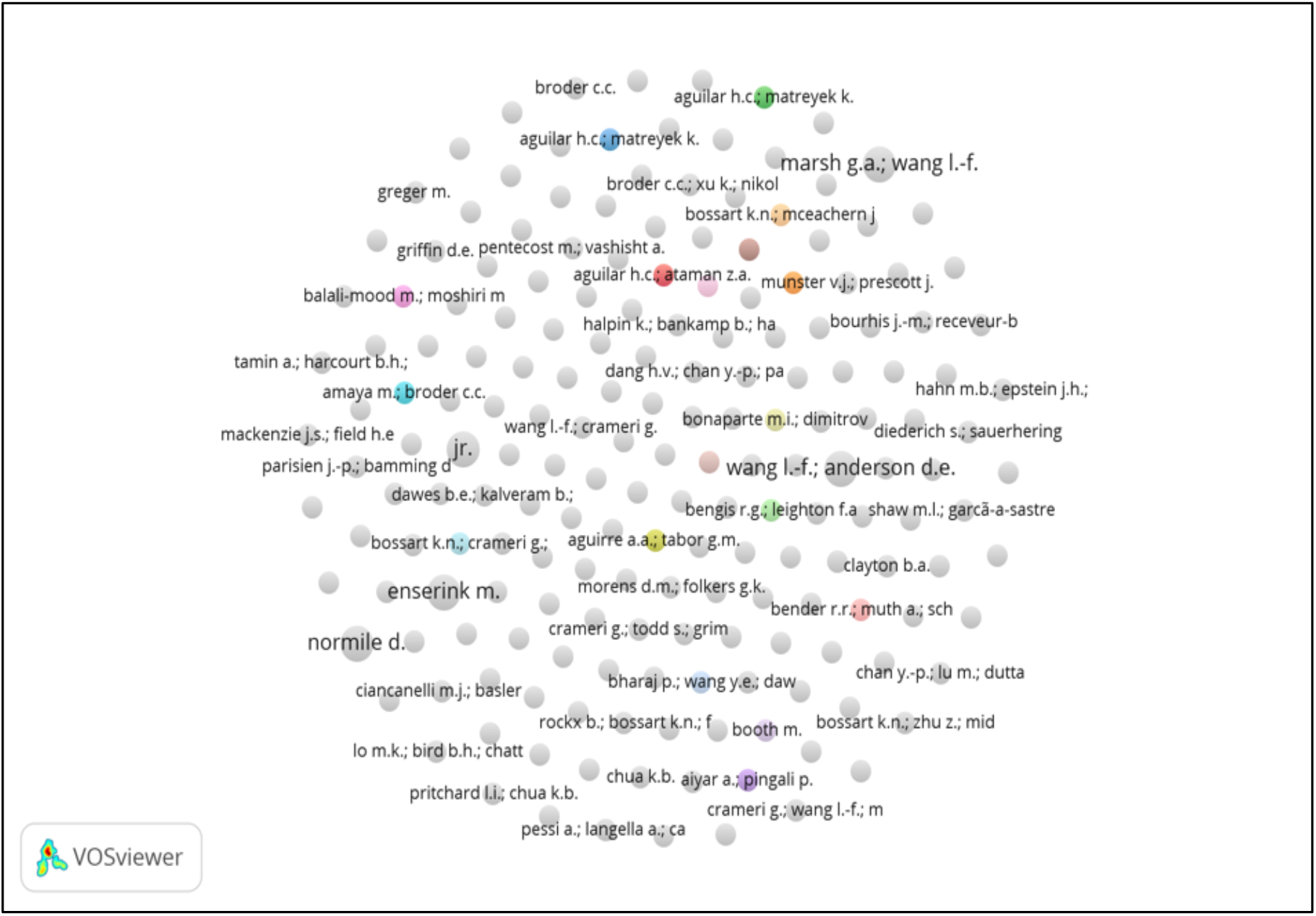
Co-authorship exploration by authors.

#### 10. Co-occurrence exploration of keywords

Figure 4 illustrates the interconnected nature of keywords, where we used the default threshold of minimum 5 occurrences per keyword. The co-occurrence analysis was conducted on 1000 keywords out of 1060 keywords that met the default threshold, providing a comprehensive view of the relationships and frequency of use among them. The key word Nipah Virus (571 occurrence) has the highest number of occurrence and it also possess the highest total link strength value with 17719. Human holds the second position with 533 occurrences, followed by nonhuman (511). This approach highlights the significant terms and their interconnections within the research domain, offering valuable insights into prevalent themes and trends.

**Figure 4:**
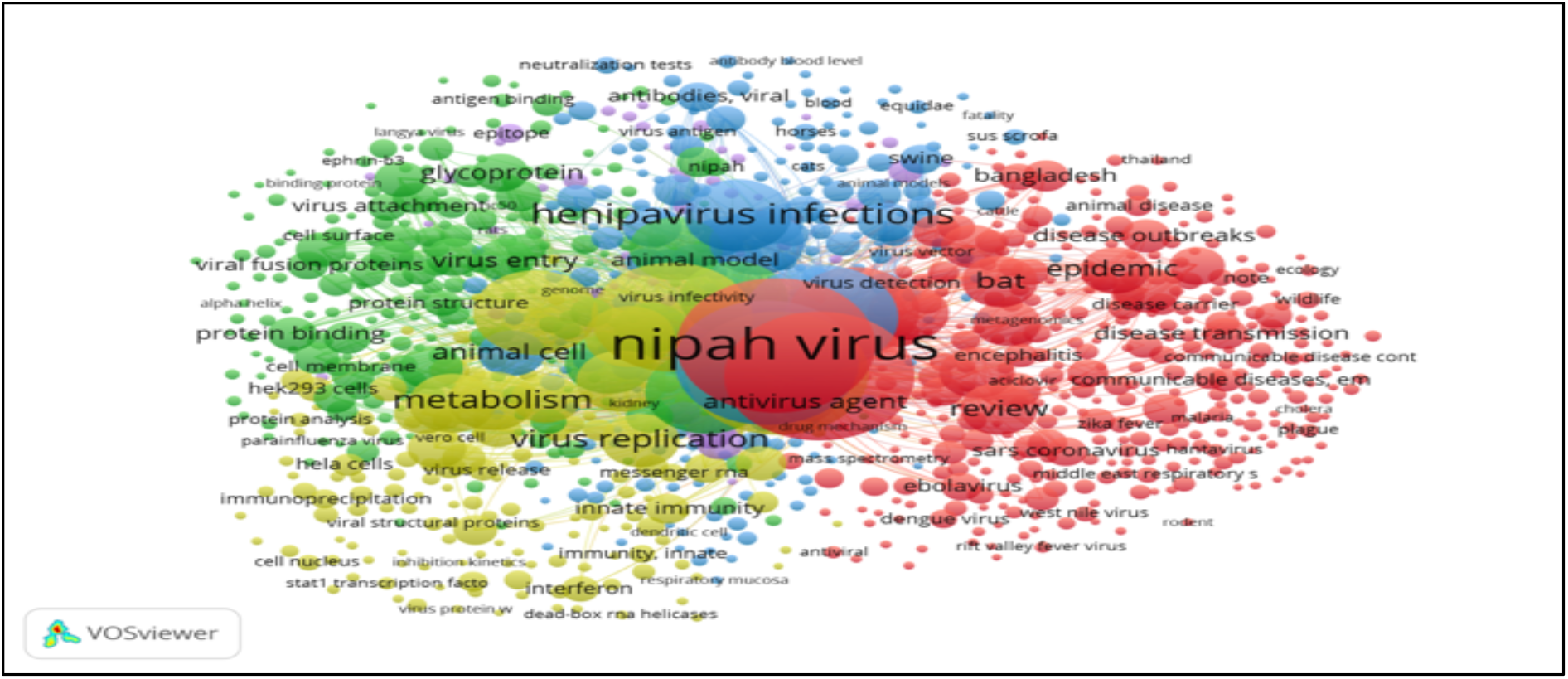
Co-occurrence exploration by keywords.

#### 11. Co-citation exploration for cited authors

Co-citation exploration for authors has been analysed by VoS Viewer software by setting a threshold of atleast 50 citations per author and is depicted in figure 5. Among 56738 authors, 398 met this threshold and were included for network analysis.

The analysis revealed that Ksiazek t.g. has emerged as highest co-cited author accumulating a total of 197186 link strengths along with 925 citations, followed by Broder C.C and Wang lin.fa. This analysis underscores the pivotal contributors and their collaborative impact within the research community.

**Figure 5:**
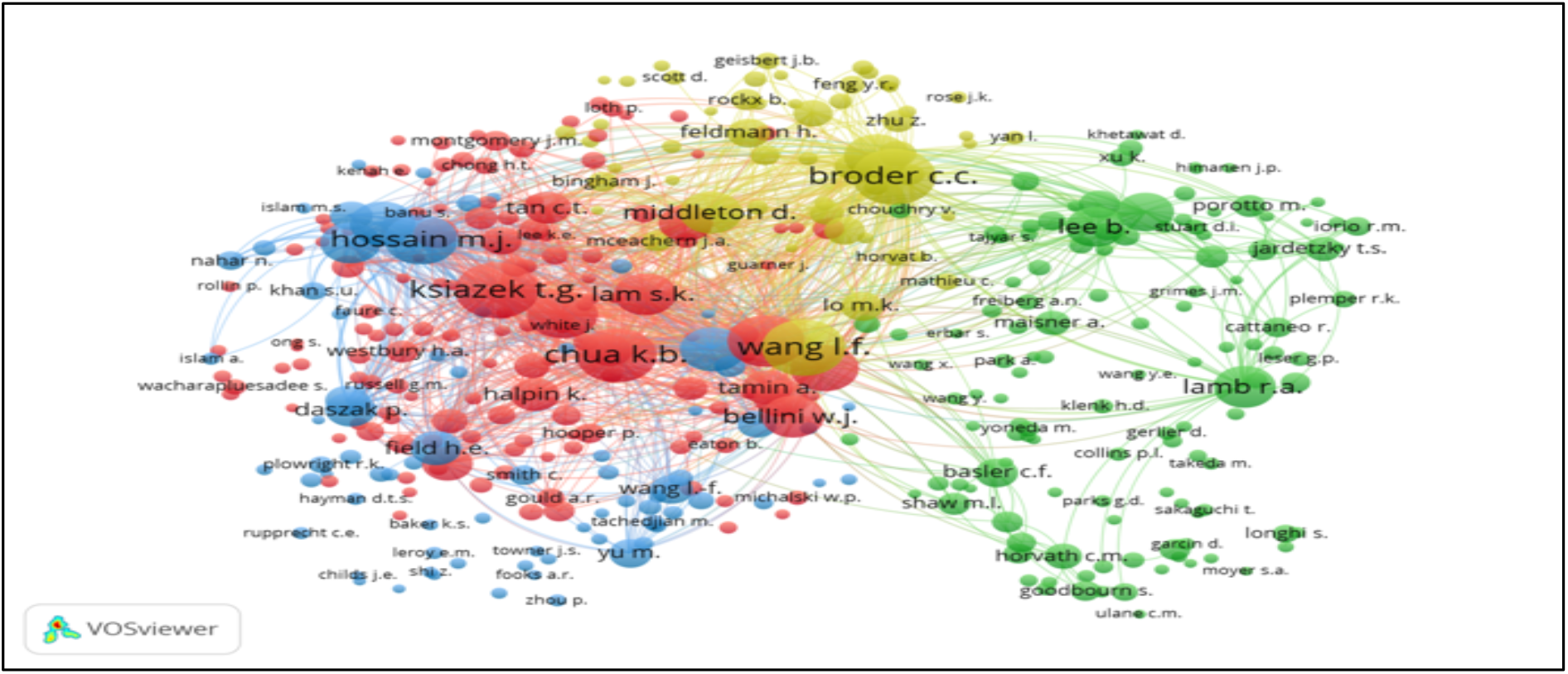
Co-citation exploration by authors.

#### 12. Co-citation exploration for cited references

The co-citation network exploration among the cited references is shown in figure 6. We have used a specific threshold of 10 citations per cited reference. Among 34215 cited references, 82 meet this threshold. The reference “negrete o.a., et al., ephrinb2 is the entry receptor for nipah virus, an emergent deadly paramyxovirus, nature, 436, pp. 401-405, (2005)” emerged top with highest citation (51) with link strength of 285. Following this, chua k.b., bellini w.j., rota p.a., harcourt b.h., tamin a., et al., nipah virus: a recently emergent deadly paramyxovirus, science, 288, pp. 1432-1435, (2000), received (48) citations with link strength of 293.

**Figure 6:**
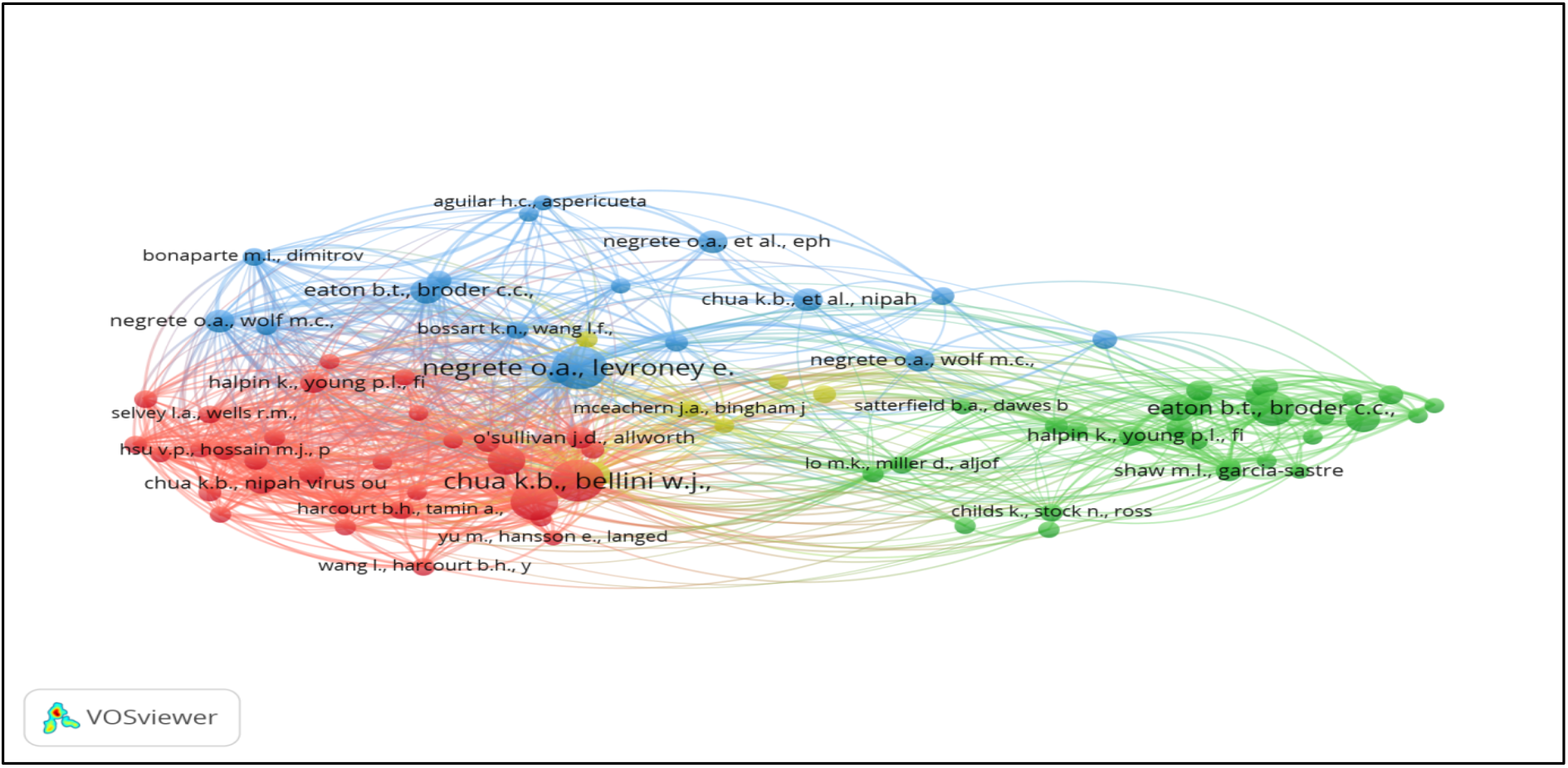
Co-citation exploration by reference.

The analysis underscores the critical works shaping the discourse in nipah virus research, highlighting the most influential studies within the co-citation network.

## Discussion

We analyzed 725 publications focusing on nipah virus, published between 2000 and 2025; this amounts to 96.15% of the total research on nipah virus published on Scopus database. Initially we conducted a numerical examination of the information and then VoS Viewer software having latest version (1.6.18) was used for the network visualization. In order to ascertain the output categorized by the years, countries, fields of study and document types. The first report on the Nipah virus as academic publication came in 2000. Thereafter, the number of publications saw a gradual increase, reaching the highest peak of 56 publications in 2020 and 2023. In 2025, the interest has been resurgent with 8 publications recorded so far and more expected to follow.

Countries such as USA, India, Australia, and United Kingdom have produced maximum publications. By doing the qualitative analysis, the most cited countries are the USA (54.6%), followed by Australia (17.9%) and India (10%) respectively. A comparison of these results shows that USA has published maximum documents on nipah virus research and thus becomes the top influential country followed by Australia and India, which makes their position in top 3 influential country lists.

Among all the disciplines that have been taken into consideration, Immunology & Microbiology (N=345) secures the highest position in number of publication related to nipah virus, followed by biochemistry, genetics and molecular biology (223) and agriculture and biological science (172). Apart from core science fields, various publications have came out of Arts and humanities too.

The analysis of document types revealed that majority of the publications falls in the category of research articles (479), followed by review articles (147). However, the fewer number of publications have been found in other domains of documents like book chapter (56), note (24), editorial (18) etc.

While analyzing the influential authors and documents in this field of study, Wang lin-fa et al. was found to be most influential author with 4437 citations, followed byBroder C. et al.(3079) and Lee behnur et. al (2298). These highly cited works demonstrates the significant contributions and ongoing relevance of key researchers in advancing the nipah virus research.

While analyzing the journal citations, we found that journal of virology has highest number of documents (111) as well as the citation score (7527), followed by science and nature journal having a citation score of (2387) and (2333) respectively. For Q-index analysis we found that 8 journals out of top 10 were having a Q1 quartile value. The h-index analysis revealed that Nature journal has the highest h-index value of 1391 followed by Science (1336). These insights underline the significant scholarly interest and impactful research being conducted in the field of virology.

International collaborations within the subject field revealed a fascinating pattern through co-authorship network analysis among researchers from different research institutes and different countries. The results revealed that the Department of pathology, University of Texas, USA has the biggest research network connecting 12 institutions, followed by Department of microbiology and immunology, Uniformed Services University, USA. Also, we found that the United States collaborated with maximum countries followed by Australia and India respectively.

The data obtained from analysis of co-occurrence of keywords revealed that the key word “Nipah Virus” appears most frequently with 571 occurrence, followed by “human” (533) and nonhuman (511). This analysis of keyword trends provides valuable insights into the current research priorities and potential directions for future studies.

The co-citation network analysis revealed that among 56738 authors, Ksiazek t.g. et. al. secured the highest number of citations. He also boasts the highest link strength of 197186 underscoring his significant influence and critical role in research landscape of Nipah virus.

## Conclusion

This analysis underscores the essential contributions shaping the academic discourse in Nipah virus research, bringing to light the most influential studies within the co-citation and co-occurrence network analysis. By identifying these keywords, we gain valuable insights into the foundational research driving advancements in this research area. The recognition of influential studies not only highlights the depth and breadth of scholarly engagement but also provides a roadmap for future research endeavours. The detailed mapping of the co-citation network further illuminates the interconnectedness of various research themes and the collaborative efforts that boost the field of Nipah virus research in different horizons.

## Data Availability

All data produced in the present study are available upon reasonable request to the authors.

## Acknowledgement

- Indian Council of Medical Research
- KSCSTE

